# Efficacy and safety of convalescent plasma and intravenous immunoglobulin in critically ill COVID-19 patients. A controlled clinical trial

**DOI:** 10.1101/2021.03.28.21254507

**Authors:** Jose Lenin Beltran Gonzalez, Mario González Gámez, Emmanuel Antonio Mendoza Enciso, Ramiro Josue Esparza Maldonado, Daniel Hernández Palacios, Samuel Dueñas Campos, Itzel Ovalle Robles, Mariana Jocelyn Macías Guzmán, Andrea Lucia García Díaz, César Mauricio Gutiérrez Peña, Ana Lilian Reza-Escalera, Maria Teresa Tiscareño Gutierrez, Elva Galvan-Guerra, Maria del Rocio Dorantes Morales, Lucila Martínez Medina, Victor Antonio Monroy Colin, Arreola Guerra Jose Manuel

**Affiliations:** Internal Medicine Department Centenario Hospital Miguel Hidalgo; Laboratorio Clinico del Campestre; Aguascalientes State Transfusion Center; Pediatrics Department Centenario Hospital Miguel Hidalgo

## Abstract

**Background:** The proportion of critically ill COVID-19 patients has collapsed hospital care worldwide. The need for alternative therapies for this group of patients is imperative. This study aims to compare the safety and efficacy of convalescent plasma (CP) compared with human immunoglobulin (IVIg) in patients requiring the administration of high oxygen levels or mechanical ventilation.

**Methods:** This is a controlled, randomized, open clinical trial of patients with pneumonia secondary to SARS-CoV-2 infection, that fulfilled criteria for severe or critical disease. They were randomized in a 1:2 ratio; group 1 was administered IVIg at a dose of 0.3 grams per kilogram of ideal weight, in an 8-hour infusion every 24 hours, for 5 days. Group 2 was administered 200 ml of CP infused in 2 hours, for 2 days. The primary outcomes were duration of hospitalization and mortality at 28 days.

**Results:** One hundred and ninety (190) patients were randomized; 130 to the CP group, and 60 to the IVIg group. Their average age was 58 years (IQR 47 – 72), and most were male (n= 119, 62.6 %). On inclusion, 85.2 % of patients (n=162) were on invasive mechanical ventilation therapy. Overall mortality in all included patients was 53 % (n= 102), with a median follow-up of 14 days (IQI 8 – 26). Mortality at 28 days was 45.2 % (n=86). In the intention-to-treat analysis, there was no difference between groups neither in mortality on follow-up (53.8 vs. 53.3, p =1.0) nor at 28 days (46.2 vs 43 %, p=0.75, Log Rank p = 0.83). Per-protocol analysis between treatment groups revealed no difference in mortality throughout hospitalization (51.5 vs 51.4 %, p=1.0) nor after 28 days (42.1 vs 42.87 %, p=0.92 Log Rank p = 0.54). Only 23 patients in the CP group received plasma with detectable anti-SARS-CoV-2 antibodies.

**Conclusions:** In critically ill patients or on invasive mechanical ventilation for treatment of Covid-19, the use of CP is not superior to IVIg in terms of hospitalization duration or mortality. The use of CP is based on complex logistics and requires an assured level of antibodies if used therapeutically. IVIg does not appear to be useful in this group of patients.

**clinicaltrials.gov identifier: NCT04381858**.

## INTRODUCTION

A new class of coronavirus was reported in December 2019 as a cause of the acute respiratory syndrome (SARS-CoV-2) in Wuhan China, leading to the development of COVID-19 disease, a public health emergency that has affected most of the world’s population and has led to thousands of deaths (1,2); in the United States, this disease is among the leading causes of death among adults above the age of 45 since October 2020 (3).

COVID-19 manifestations may vary from mild symptoms to the development of the Acute Respiratory Distress Syndrome (4), for which therapeutic strategies have, to date, been limited. Multiple treatment options have been used but only a few drugs have been able to decrease mortality in patients with pulmonary disease; their usefulness has varied depending on disease severity and in some cases, their availability has been limited due to the magnitude of the affected population throughout the world (5-8).

Interest in the use of convalescent plasma (CP) arose since the beginning of the pandemic, based on the awareness of passive immunization strategies previously used in other viral diseases (9-11); it was considered a safe and low-cost option so on August 23, 2020, the Food and Drug Administration (FDA) in the United States authorized its emergency use, although the development of controlled clinical trials was still necessary to prove its efficacy (12).

The use of human immunoglobulin (IVIg) has been proposed as therapy of viral diseases such as influenza A H1N1. In a Covid-19 retrospective study, its administration in the first 48 hours after symptom development decreased mortality and the risk of complications (13,14). However, there is still great uncertainty on its possible effects due to the paucity of studies designed to test its efficacy.

This trial analyzes the safety and efficacy of the administration of CP in comparison with IVIg in critically ill patients due to SARS-CoV-2 infection.

## MATERIAL AND METHODS

This is a controlled, randomized, open clinical trial including patients with secondary pneumonia to SARS-CoV-2 infection, and fulfilling severe or critical disease criteria, to compare the efficacy and safety of CP administration in comparison with the use of IVIg.

We obtained convalescent plasma from individuals with a reactive SARS-CoV-2 nasopharyngeal swab, a second negative test, and in an asymptomatic state in the previous 14 days. In the absence of a second RT-PCR, we included donors with an initially positive test, a minimum disease course of 28 days, and that remained asymptomatic during the 14 days prior to donation.

Patients that were plasma donation candidates were selected in accordance with Mexican Official Norms (15). Serologies for human immunodeficiency virus, hepatitis B, hepatitis C, trypanosomiasis, and syphilis were also obtained.

Convalescent plasma was obtained by puncture of a peripheral vein and plasmapheresis (TERUMO BCT, model Trima Accel^®^). The quantity of extracted plasma varied according to body weight between 200 ml and 600 ml, which were divided into 200 ml units, and subsequently stored and frozen at -70°C.

Serum aliquots from donor patients were safeguarded to document the presence of anti-SARS-CoV-2 IgG antibodies by immunochemiluminescence (ARCHITECT ABBOTT). Given the study’s temporality and the availability of serological tests, all serological results were obtained after plasma administration.

Patients included in the study had to fulfill the operational definition of a suspected or confirmed case of COVID-19, and present criteria of severe pneumonia according to the ATS/IDSA guidelines (16). We considered for inclusion those patients with: 1) A positive nasopharyngeal and oropharyngeal swab RT-PCR for SARS-CoV-2, 2) Pneumonia diagnosed by high-resolution CT scan of the chest, and a pattern suggesting coronavirus infection, 3) Recently developed hypoxemic respiratory failure or acute clinical exacerbation of pre-existing pulmonary or heart disease, 4) Requirement of respiratory support with a high-flow nasal cannula, defined as 60 liters with a 90% inspired oxygen fraction or invasive mechanical ventilation with an orotracheal tube.

All hospitalized patients received pharmacological thromboprophylaxis with low-weight molecular heparin or unfractionated heparin, according to international guidelines (17,18).

On the last week in June and based on the evidence obtained in the RECOVERY trial, we began the administration of dexamethasone, 6 mg intravenously, every 24 hours for 10 days (19).

Patients were administered ivermectin, 12 mg if their weight was below 80 Kg, and 18 mg if it was above 80 Kg; this treatment was established in all patients due to its theoretical and potential therapeutic benefit. We concomitantly conducted a clinical trial with patients requiring hospitalization but that were not critically ill, to compare the safety and efficacy of ivermectin and hydroxychloroquine (Clinical Trials Identifier: NCT04391127).

We performed an intermediate analysis in August 2020, that proved the therapeutic futility of ivermectin and hydroxychloroquine, so their administration was withdrawn in protocol patients.

Patients that were included signed an informed consent form, either personally or if unable due to illness severity, by the responsible relative whose authorization was requested by telephone following hospital safety measures.

Two treatment groups were randomized in a 1:2 ratio. Group 1 was administered human immunoglobulin at a dose of 0.3 grams per kilogram of ideal body weight, in an 8-hour infusion every 24 hours, for 5 days. Group 2 was administered 200 ml of CP over 2 hours, every 24 hours, for 2 days.

On admission, we obtained blood samples to determine arterial blood gases, a complete blood count, blood chemistry, and prognostic markers such as fibrinogen, D-dimer, ferritin, troponin I, C-reactive protein, prothrombin time, and activated thromboplastin time; follow-up laboratory tests were obtained according to the treating physician’s criterion.

The primary outcome was established as the duration of hospitalization, and the safety outcome was patient death and/or the development of adverse effects following treatment administration. The secondary outcome was the duration of invasive mechanical ventilation.

This study was approved by the ethics committee of the Centenario Hospital Miguel Hidalgo, on April 15, 2020, with the identifier number 2020-R-25. It was also registered in the clinicaltrials.gov site with the identifier NCT04381858.

## Statistical Analysis

Descriptive statistics were used depending on the measurement level. The distribution of continuous variables was evaluated with the Kolmogorov Smirnov test. Continuous variables with a normal distribution were expressed as means and standard deviation, while variables with an abnormal distribution were expressed as medians and interquartile intervals. Categorical variables were expressed as relative and absolute frequencies. Continuous variables in between-group analyses were evaluated according to their distribution, with Student’s T or Mann-Whitney’s U. Dichotomic and ordinal variables were analyzed with the χ^2^ test or Fisher’s exact test, as needed. Survival analysis for the death outcome was conducted with Kaplan Meier curves, and between-group comparisons were obtained with the Log-rank test. A p-value below 0.05 was considered significant. We used Microsoft Excel 2013 and STATA version 11.1 software.

## RESULTS

Between May 5 and October 17, 2020, we recruited 193 patients, 3 of whom were eliminated due to hospital transfer and the inability to continue their treatment and surveillance. In the end, 190 patients were randomized: 130 to the CP group and 60 to the IVIg group. (Figure 1)

**Figure 1.**
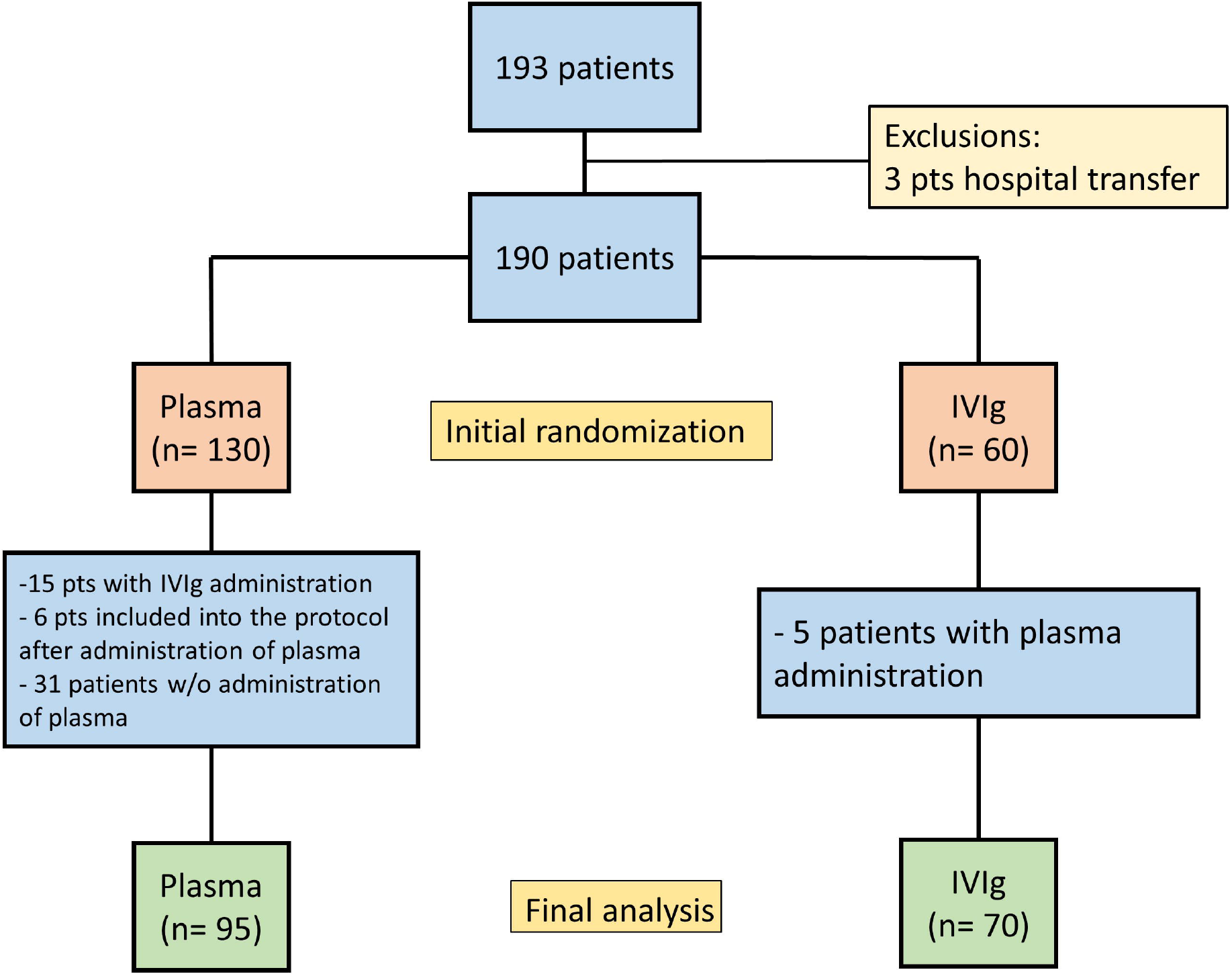
Recruited Patients. Among the 193 recruited patients, 3 were excluded due to hospital transfer. In the group randomized to CP, 15 patients were administered IVIg due to the unavailability of plasma, 6 patients received CP without randomization but had signed the informed consent form, 31 patients did not receive plasma due to its unavailability. In the IVIg group, 5 patients were transferred to the CP group due to IVIg unavailability. Per-protocol, 95 patients received CP and 70 received IVIg.

Average age was 58 years (IQR 47 – 72), with a predominance of males (n= 119, 62.6 %). The most frequent comorbidity was obesity/overweight (n = 153, 80.5 %), followed by systemic arterial hypertension (n = 67, 35.2 %), and diabetes mellitus (n= 66, 34.7 %). (Table 1 and Addendum 1)

**Table 1.** Demographic characteristics, population comorbidities, and additional drugs administered during hospitalization (Intention-to-treat analysis).

| Variable | All (n=190) | Plasma (n=130) | IVIg (n=60) | P val |
| --- | --- | --- | --- | --- |
| Sex Male, n (%) | 119 (62.6) | 80 (61.5) | 39 (65) | 0.74 |
| Age, m (S) | 58 (47 – 72) | 60 (48 - 74) | 55 (46.5 - 67) | 0.15 |
| Comorbidities, n (%) |  |  |  |  |
| Diabetes | 66 (34.7) | 42 (32.3) | 24 (40) | 0.30 |
| SAH | 67 (35.2) | 46 (35.3) | 21 (35) | 0.95 |
| Overweight | 74 (38.9) | 55 (42.3) | 19 (31.6) | 0.2 |
| Obesity | 79 (41.5) | 53 (40.7) | 26 (43.3) | 0.75 |
| Stroke | 2 (1.05) | 2 (1.54) | 0 | 1.0 |
| Smoker | 78 (41) | 55 (42) | 24 (40) | 0.77 |
| Alcohol /Drugs | 6 (3.1) | 3 (2.3) | 3(5) | 0.38 |
| Heart disease | 6 (3.1) | 4 (3.1) | 2 (3.3) | 1.0 |
| Pulmonary disease | 9 (4.7) | 6 (4.6) | 3 (5.0) | 1.0 |
| CKD | 6 (3.1) | 4 (3.1) | 2 (3.3) | 1.0 |
| Hypothyroidism | 4 (2.1) | 3 (2.3) | 1 (1.7) | 1.0 |
| HIV | 1 (0.53) | 0 | 1 (1.7) | 0.31 |
| Cancer | 1 (0.53) | 0 | 1 (1.7) | 0.31 |
| Transplant | 1 (0.53) | 1 (0.7) | 0 | 1.0 |
| Autoimmunity | 2 (1.05) | 1 (0.7) | 1 (1.7) | 0.53 |
| BMI, med (IQR) | 28.4 (25.3 – 32.7) | 28.3 (25.4 – 33.2) | 28.9 (24.9 – 32.7) | 0.62 |
| **Other drugs** |  |  |  |  |
| Antibiotics | 107 (56.3) | 75 (57.6) | 32 (53.3) | 0.63 |
| Carbapenem drugs | 76 (40) | 50 (38.4) | 16 (43.3) | 0.52 |
| PBI | 187 (98.4) | 127 (97.6) | 60 (100) | 0.23 |
| Dexamethasone | 157 (82.6) | 109 (83.8) | 48 (80) | 0.54 |
| Ivermectin | 116 (61) | 76 (58.4) | 40 (66.6) | 0.28 |

At inclusion, 85.2 % of patients (n=162) required invasive mechanical ventilation, and the remaining patients were managed with high-oxygen flow devices. The average initial oxygenation index was 165 (IQI 96 – 240) (Table 2). Prognostic clinical markers on admission revealed a median APACHE II score of 12 points (IQI 9 – 16), SOFA 3 points (IQI 2 – 4), and CURB-65, 2 points (IQI 1 – 2) (Table 3). Among inflammation biomarkers, median LDH was 479 U/L (IQI 367 – 658), D-dimer was 1,300 ng/mL (IQI 800 – 3000), and ferritin was 584 ng/mL (IQI 324 – 1080). (Table 4)

**Table 2.**
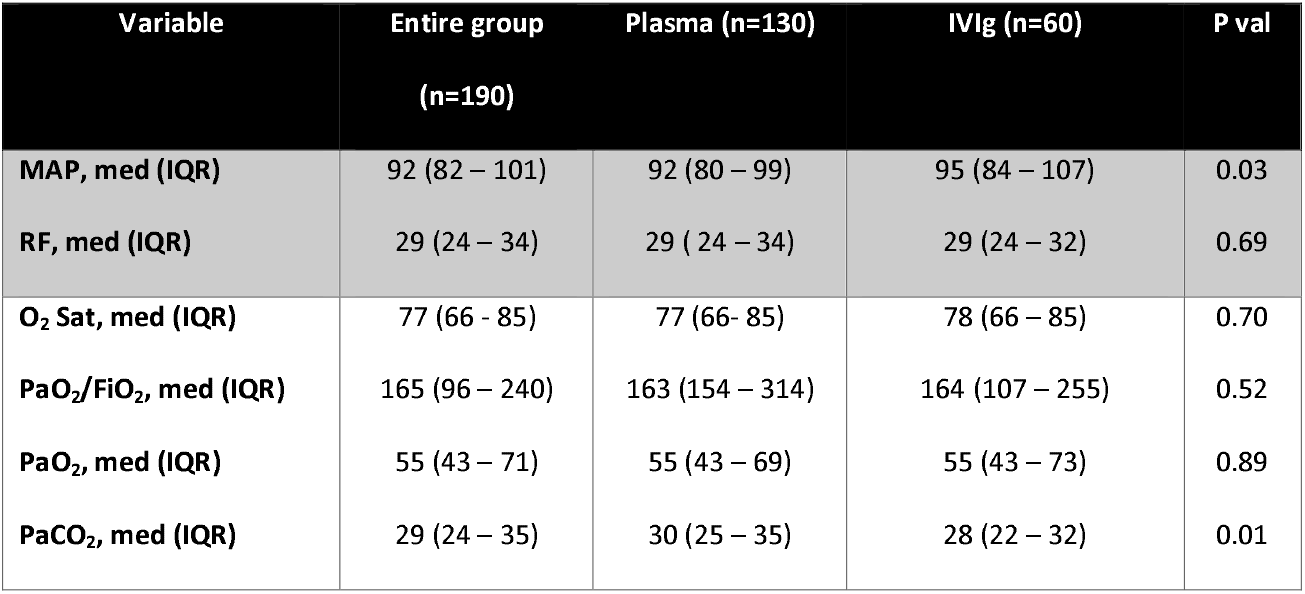
Vital signs and oxygenation index on admission (Intention-to-treat analysis).

| **Variable** | **Entire group**<br>**(n=190)** | **Plasma (n=130)** | **IVIg (n=60)** | **P val** |
| --- | --- | --- | --- | --- |
| **MAP, med (IQR)** | 92 (82 – 101) | 92 (80 – 99) | 95 (84 – 107) | 0.03 |
| **RF, med (IQR)** | 29 (24 – 34) | 29 ( 24 – 34) | 29 (24 – 32) | 0.69 |
| **O<sub>2</sub> Sat, med (IQR)** | 77 (66 - 85) | 77 (66- 85) | 78 (66 – 85) | 0.70 |
| **PaO<sub>2</sub>/FiO<sub>2</sub>, med (IQR)** | 165 (96 – 240) | 163 (154 – 314) | 164 (107 – 255) | 0.52 |
| **PaO<sub>2</sub>, med (IQR)** | 55 (43 – 71) | 55 (43 – 69) | 55 (43 – 73) | 0.89 |
| **PaCO<sub>2</sub>, med (IQR)** | 29 (24 – 35) | 30 (25 – 35) | 28 (22 – 32) | 0.01 |

**Table 3.** Prognostic Scales (Intention-to-treat analysis).

**Addendum 2.** Vital signs and oxygenation index on admission (Per-protocol analysis).
| **Variable** | **All patients (n=190)** | **Plasma (n=130)** | **IVIg (n=60)** | **P val** |
| --- | --- | --- | --- | --- |
| **SOFA med (IQR)** | 3 (2 – 4) | 3 (2 – 4) | 3 (2 – 4) | 0.97 |
| **SOFA > 2, n (%)** | 136 (71.3) | 93 (72) | 43 (71.6) | 0.95 |
| **APACHE II, med (IQR)** | 12 (9 – 16) | 12 (9 – 16) | 12 (9 – 17) | 0.77 |
| **APACHE > 8, n (%)** | 161 (85) | 109 (84) | 52 (86.6) | 0.69 |
| **CURB-65, med (IQR)** | 2 (1 – 2) | 2 (1 – 3) | 1 (1 – 2) | 0.41 |
| **CURB-65 ≥2, N (%)** | 98 (51.8) | 70 (54) | 28 (46.6) | 0.33 |

**Table 4.** Inflammation markers (Intention-to-treat analysis).

**Addendum 3.** Prognostic Scales (Per-protocol analysis).
| Variable, med (IQR) | All patients (n=190) | Plasma (n=130) | IVIg (n=60) | P val. |
| --- | --- | --- | --- | --- |
| Lactate | 1.6 (1.1 – 2) | 1.6 (1.1 – 2) | 1.45 (1.2 – 2) | 0.82 |
| Creatinine | 0.9 (0.7 – 1.4) | 0.9 (0.7 – 1.4) | 0.9 (0.7 – 1.4) | 0.87 |
| LDH | 479 (367 – 658) | 483 (362 – 660) | 482 (376 – 658) | 0.76 |
| Troponin | 0.021 (0.012 – 0.11) | 0.021 (0.012 – 0.12) | 0.015 (0.11– 0.09) | 0.56 |
| D-Dimer | 1.3 (0.8 – 3.0) | 1.3 (1.2 -3.3) | 1.4 (0.8 – 2.4) | 0.70 |
| Ferritin | 584 (324 – 1080) | 562 (308 – 1040) | 636 (358 – 1170) | 0.21 |
| Hb | 14.4 (13 – 15.5) | 14.2 (12.9 – 15.4) | 14.8 (13.6 – 15.7) | 0.08 |
| Leukocytes | 11.8 (8.4 – 15.2) | 11.7 (8.6 – 14.6) | 11.9 (7.5 – 15.8) | 0.89 |
| Neutrophils | 9.9 (6.9 – 13.2) | 9.8 (7.1 – 13) | 10.4 (6.2 – 13.3) | 0.93 |
| Lymphocytes | 1.02 (0.7 – 1.5) | 1.02 (0.7 – 1.5) | 0.9 (0.69 – 1.52) | 0.84 |
| CRP 1 | 192 (56 -270) | 195 (55 – 270) | 205 (66 – 300) | 0.67 |
| CRP 2 | 163 (60 – 232) | 194 (62 – 249) | 86 (77 – 175) | 0.23 |
| CRP 3 | 170 (47 – 231) | 189 (60 – 250) | 130 (38 – 204) | 0.40 |
| Ferritin 1 | 584 (402 – 1080) | 562 (308 – 1040) | 636 (358 – 1170) | 0.21 |
| Ferritin 2 | 710 (402 – 1280) | 712 (389 – 1280) | 693 (446 – 1390) | 0.50 |
| Ferritin 3 | 622 (342 – 1035) | 674 (337 – 1070) | 659 (454 – 993) | 0.93 |
| D-Dimer 1 | 1.3 (0.8 – 2.9) | 1.3 (0.8 – 3.8) | 1.4 (0.8 – 2.4) | 0.87 |
| D-Dimer 2 | 1.8 (1.1 – 4.3) | 1.6 (1.1 – 5.1) | 2.1 (1.2 – 3.6) | 0.71 |
| D-Dimer 3 | 1.8 (0.9 – 2.5) | 1.8 (1.08 – 2.6) | 1.7 (0.6 – 2.4) | 0.34 |

Analysis of between-group differences only revealed that systemic arterial pressure was lower in the CP group vs IVG (92 vs 95 mm Hg, p=0.03), and PaCO2 was lower in the IVIg group (30 vs 28 mmHg, p=0.01). There were no significant differences in the remaining characteristics.

In terms of protocol analysis, 95 patients received at least one plasma unit, and 70 patients received IVIg. There were no significant differences between both groups. (Addenda 2 – 5)

### Outcomes

Mortality among included patients was 53 % (n= 102) with a median follow-up of 14 days (IQI 8 – 26). Mortality at 28 days was 45.2 % (n=86). Intention-to-treat analysis showed no differences between groups or in mortality throughout follow-up (53.8 vs 53.3, p =1.0) or at 28 days (46.2 vs 43 %, p=0.75). (Table 5 and Figure 2)

**Table 5.**
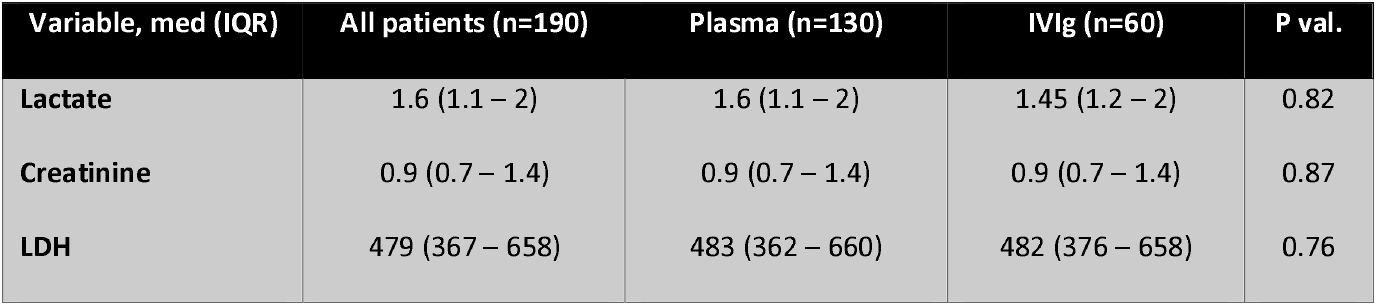
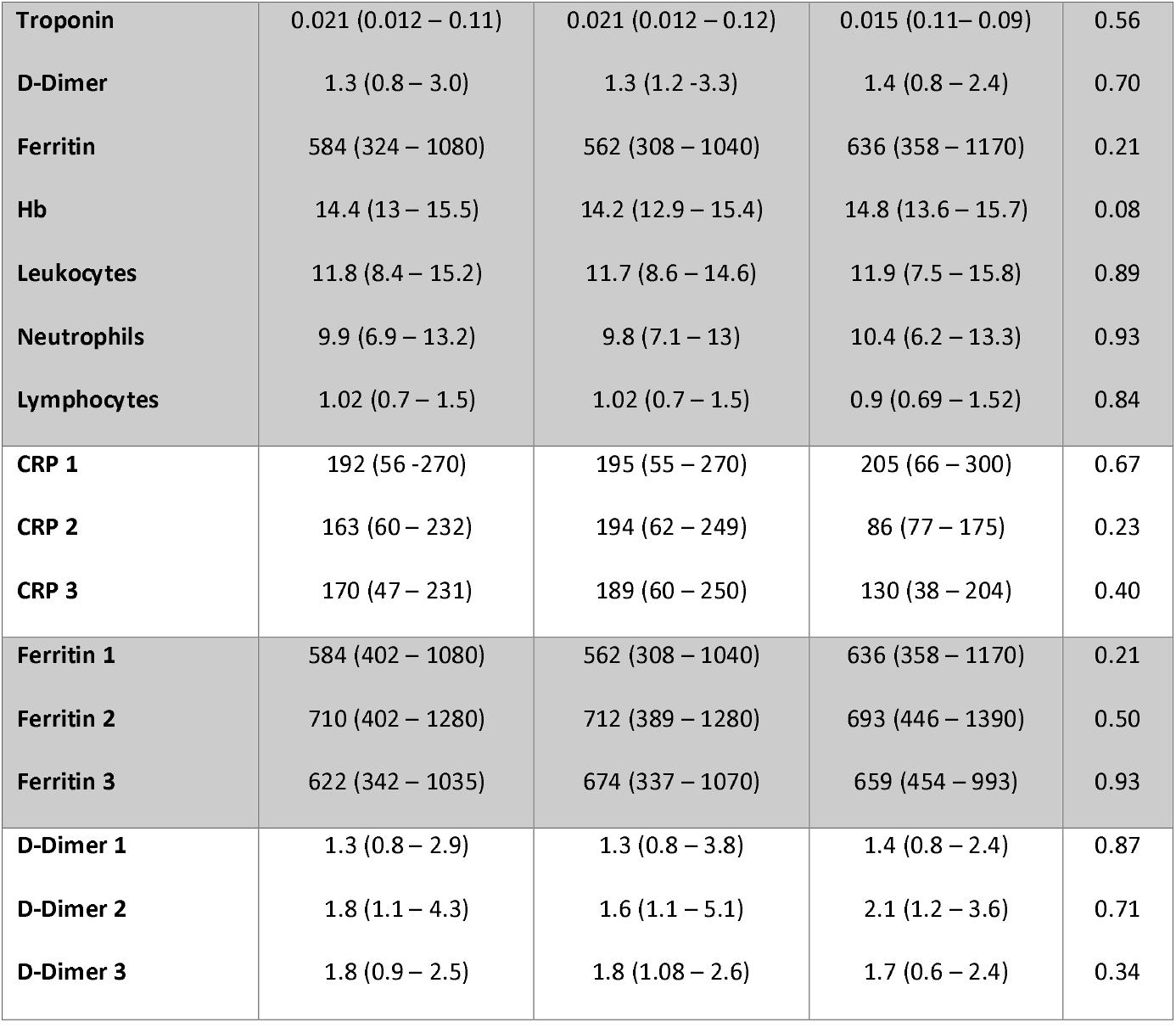
Outcomes (Intention-to-treat analysis).

**Addendum 4.** Inflammation markers (Per-protocol analysis).
| **Variable, med (IQR)** | **All patients (n=190)** | **Plasma (n=130)** | **IVIg (n=60)** | **P val.** |
| --- | --- | --- | --- | --- |
| **Lactate** | 1.6 (1.1 – 2) | 1.6 (1.1 – 2) | 1.45 (1.2 – 2) | 0.82 |
| **Creatinine** | 0.9 (0.7 – 1.4) | 0.9 (0.7 – 1.4) | 0.9 (0.7 – 1.4) | 0.87 |
| **LDH** | 479 (367 – 658) | 483 (362 – 660) | 482 (376 – 658) | 0.76 |
| **Troponin** | 0.021 (0.012 – 0.11) | 0.021 (0.012 – 0.12) | 0.015 (0.11– 0.09) | 0.56 |
| **D-Dimer** | 1.3 (0.8 – 3.0) | 1.3 (1.2 -3.3) | 1.4 (0.8 – 2.4) | 0.70 |
| **Ferritin** | 584 (324 – 1080) | 562 (308 – 1040) | 636 (358 – 1170) | 0.21 |
| **Hb** | 14.4 (13 – 15.5) | 14.2 (12.9 – 15.4) | 14.8 (13.6 – 15.7) | 0.08 |
| **Leukocytes** | 11.8 (8.4 – 15.2) | 11.7 (8.6 – 14.6) | 11.9 (7.5 – 15.8) | 0.89 |
| **Neutrophils** | 9.9 (6.9 – 13.2) | 9.8 (7.1 – 13) | 10.4 (6.2 – 13.3) | 0.93 |
| **Lymphocytes** | 1.02 (0.7 – 1.5) | 1.02 (0.7 – 1.5) | 0.9 (0.69 – 1.52) | 0.84 |
| **CRP 1** | 192 (56 -270) | 195 (55 – 270) | 205 (66 – 300) | 0.67 |
| **CRP 2** | 163 (60 – 232) | 194 (62 – 249) | 86 (77 – 175) | 0.23 |
| **CRP 3** | 170 (47 – 231) | 189 (60 – 250) | 130 (38 – 204) | 0.40 |
| **Ferritin 1** | 584 (402 – 1080) | 562 (308 – 1040) | 636 (358 – 1170) | 0.21 |
| **Ferritin 2** | 710 (402 – 1280) | 712 (389 – 1280) | 693 (446 – 1390) | 0.50 |
| **Ferritin 3** | 622 (342 – 1035) | 674 (337 – 1070) | 659 (454 – 993) | 0.93 |
| **D-Dimer 1** | 1.3 (0.8 – 2.9) | 1.3 (0.8 – 3.8) | 1.4 (0.8 – 2.4) | 0.87 |
| **D-Dimer 2** | 1.8 (1.1 – 4.3) | 1.6 (1.1 – 5.1) | 2.1 (1.2 – 3.6) | 0.71 |
| **D-Dimer 3** | 1.8 (0.9 – 2.5) | 1.8 (1.08 – 2.6) | 1.7 (0.6 – 2.4) | 0.34 |

**Figure 2.**
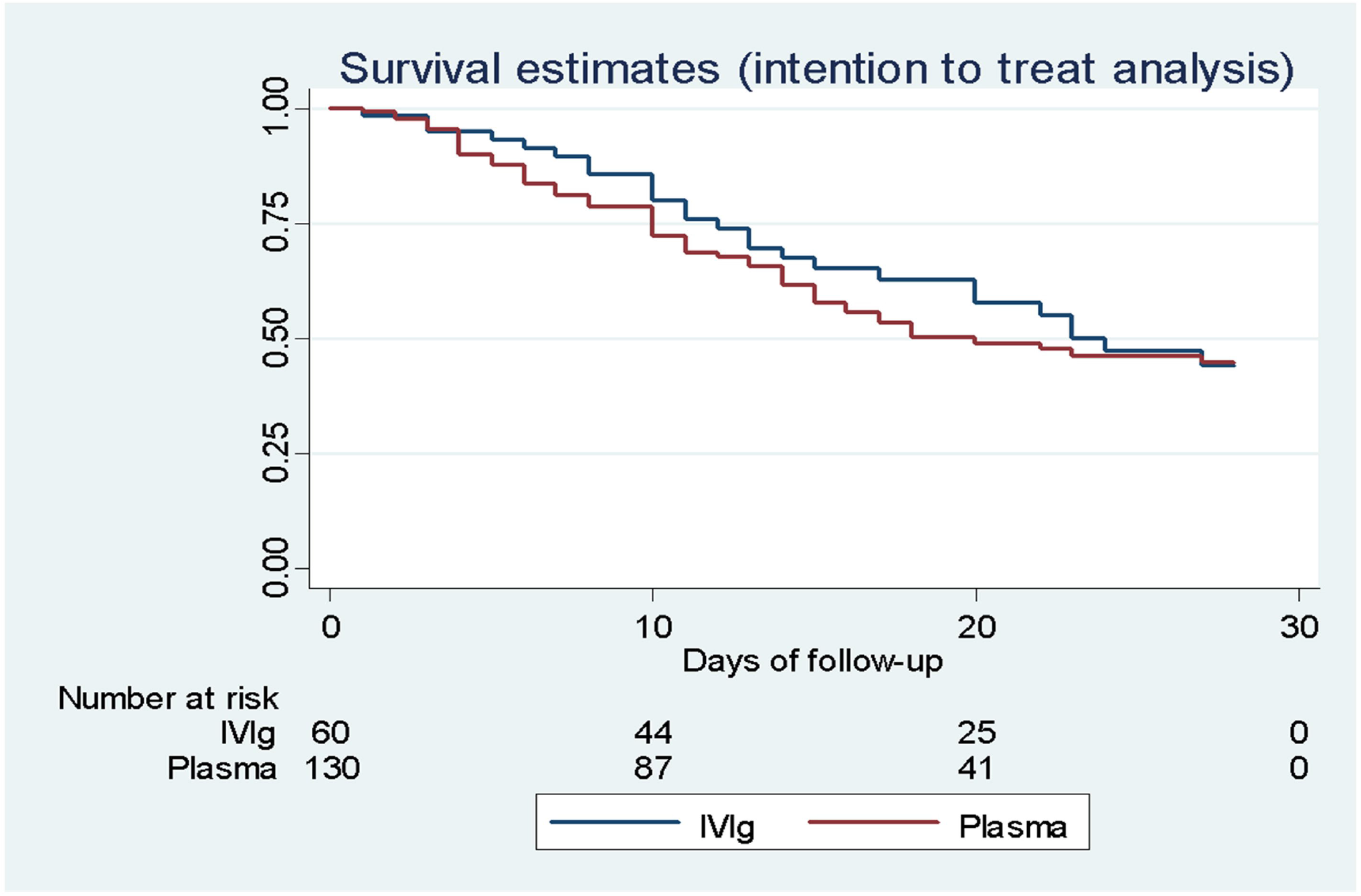
Survival at 28 days analysis. Convalescent plasma *vs* IVIg. Log Rank: p = 0.83. (Intention-to-treat analysis).

**Figure 3.**
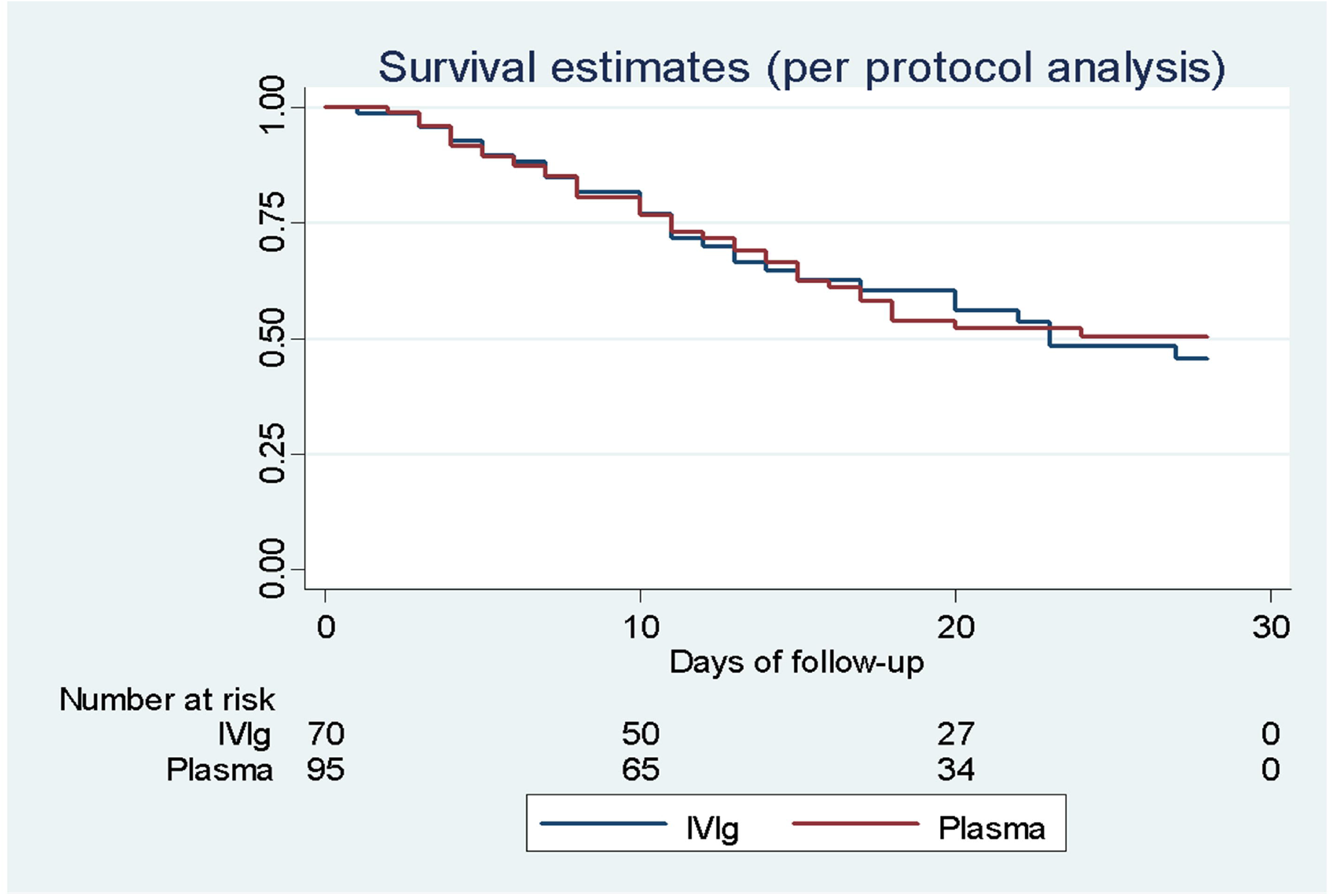
Survival at 28 days analysis. Convalescent plasma *vs* IVIg. Log Rank: p= 0.54 (Per-protocol analysis).

Per protocol analysis revealed an overall mortality of 51.5 % (n= 85), and 42.2 % at 28 days (n=70). The median duration of hospitalization was 12 days (IQR, 6 – 22). There was no difference in mortality between treatment groups throughout hospitalization (51.5 vs 51.4 %, p=1.0) or at 28 days (42.1 vs 42.87 %, p=0.92) (Table 5). The median duration of mechanical ventilation was 12 days (IQI 7 – 23), and there were no between-group differences.

Among the convalescent plasmas administered to 95 patients, the presence of anti-SARS-CoV-2 IgG antibodies was determined in only 78 cases. Twenty-three patients (29.5%) received at least one unit of plasma with antibodies. Mortality at 28-days did not differ whether antibodies were present or not (43.4 vs 40%, p= 0.81), nor did the duration of hospitalization (18 vs 13 days, p=0.08), nor the duration of mechanical ventilation (17.8 vs 18.6 days, p= 0.83.) Two plasma units were administered to 53 patients; their 28-day mortality was 33.9 %, which was not statistically different from that in the IVIg group (42.8 %, p=0.31).

Patients that did not receive treatment (n=31) had greater mortality (non-significant) in comparison with those included in the treatment groups (54.8 vs 42.4 % p= 0.20).

### Adverse Events

Two patients in the IVIg group developed adverse events associated with its administration; one had an anaphylactic reaction, and the other developed a hypertensive crisis. The IVIg infusion was immediately suspended in the first case, and antihypertensive treatment was initiated in the second case. The CP group did not develop adverse effects attributable to its administration.

The most commonly observed complication was the development of shock, which occurred in 71% of cases (n=136). Bacterial co-infections appeared in 56.6 % (n=107). No between-group differences were detected. (Addendum 5)

## Discussion

In this study, the administration of CP was not superior to the administration of IVIg, both in terms of mortality and hospitalization duration. The number of patients warranting mechanical ventilation as well as its duration was also no different between groups.

Several published clinical trials have evaluated the efficacy of CP administration in patients with COVID-19, and their results have varied in function of the group characteristics. The clinical trial by Ling Li et al was prematurely terminated due to a decrease in the number of cases during the study since only half of the initially calculated sample was recruited; they included a total of 103 patients, the primary outcome was clinical improvement at 28 days, and no significant differences were found. The subgroup of patients with severe disease had a superior outcome when compared with placebo, 91.3 vs 68.2 % (HR 2.15, IC 95% 1.07 – 4.32, p = 0.03). Further, that trial referred a significant decrease in viral load after 72 hours in the CP group, particularly in patients with life-threatening disease (20). A second trial (PLACID Trial) included 464 patients with moderate COVID-19 (PaFiO2 between 200 and 300), and there was no difference between the treatment groups in terms of disease progression or death. However, they only included 19 patients on invasive mechanical ventilation per group (21).

Simonovich et al., as part of the PlasmAr group, conducted a clinical trial with patients with severe pneumonia due to COVID-19; they found no significant clinical improvement nor differences in mortality when compared with the placebo group. Despite assuring a minimal level of anti-SARS-CoV-2 antibodies, there was no significant difference in mortality in comparison with placebo; the CP was administered after 8 days (mean) since the initiation of symptoms. That study’s inclusion criteria did not include patients on invasive mechanical ventilation. Among those that required invasive mechanical ventilation during follow-up, no difference in outcome was noted (22).

Finally, a study by Libster et al. showed decreased COVID-19 progression in elderly patients with mild infection to whom CP with high titers of anti-SARS-CoV-2 antibodies was administered within the first 72 hours from symptom onset; a dose-dependent effect was reported, according to the detected antibody titer in the administered plasma (23).

To date, there have been two clinical trials on the use of IVIg; the first included hospitalized patients with persistent hypoxemia 48 hours after admission. They included 30 patients in the IVIg group and 29 in the control group. Mortality was lower in the IVIg group (20 % vs 48.3 %, p= 0.02). (24) The second trial included patients with severe COVID-19, 52 patients were allotted to the IVIg group, and 32 to the control group. They used an IVIg dose of 400 mg/kg, for three days. No differences in mortality or duration of hospitalization were detected (25).

Our study included severely ill patients on invasive mechanical ventilation or with respiratory deterioration as a result of their high mortality and a lack of therapeutic alternatives. We compared CP against IVIg as passive immunization therapies and detected no differences in mortality or duration of mechanical ventilation, perhaps as a result of the severe disease in included cases and our inability to ascertain the level of antibodies in the CP group.

One of our study’s weaknesses is the use of convalescent plasmas in which antibodies were not measured since this is a limited resource in our setting. This occurred at the beginning of the study and later, we were only able to conduct qualitative determinations.

But this study underscores the complexity of recruiting optimal plasma donors and the logistics of its administration. This is clearly reflected in the large proportion of patients that did not receive CP. On the other hand, the IVIg group was treated at pre-established times and with established doses, with no relevant deviations from the contemplated protocol. Regardless, it did not prove to be superior to CP which suggests therapeutic futility, at least in terms of the administered dosage and the characteristics of our patient population.

## CONCLUSIONS

In gravely ill patients and those on invasive mechanical ventilation as a result of COVID-19, the use of CP without establishing an adequate level of anti-SARS-CoV-2 antibodies is not superior to the use of IVIg in terms of hospitalization duration or mortality. The use of CP is based on complex logistics and ensuring an adequate level of antibodies is pivotal to its potential therapeutic use. The administration of IVIg does not appear to be of use in this group of patients.

**Addendum 1.** Demographic characteristics, population comorbidities, and additional drugs administered during hospitalization (Per-protocol analysis).

| Variable | All patients (n=165) | Plasma (n=95) | IVIg (n=70) | P v |
| --- | --- | --- | --- | --- |
| Sex Male, n (%) | 101 (61.2) | 54 (56.8) | 47 (66.6) | 0.18 |
| Age, m (S) | 58.1 (15) | 59 (15.8) | 55 (14.3) | 0.14 |
| **Comorbidities, n (%)** |  |  |  |  |
| Diabetes | 57 (34.5) | 34 (35.4) | 23 (32.8) | 0.69 |
| SAH | 59 (35.7) | 34 (35.7) | 25 (35.7) | 0.99 |
| Overweight | 60 (36.3) | 37 (38.9) | 23 (32.8) | 0.42 |
| Obesity | 67 (40.6) | 36 (37.9) | 31 (44.3) | 0.40 |
| Stroke | 1 (0.61) | 1 (1.05) | 0 | 1.0 |
| Smoker | 66 (40) | 35 (36.8) | 31(44) | 0.36 |
| Alcohol /Drugs | 6 (3.6) | 2 (2.1) | 4 (5.7) | 0.40 |
| Heart Disease | 3 (1.8) | 1 (1.05) | 2 (2.9) | 0.57 |
| Pulmonary Disease | 10 (6) | 7 (7.4) | 3 (4.3) | 0.52 |
| CKD | 6 (3.6) | 4 (4.2) | 2 (2.9) | 1.0 |
| **Hypothyroidism** | 4 (2.4) | 3 (3.2) | 1 (1.4) | 0.63 |
| **HIV** | 1 (0.6) | 0 | 1 (1.4) | 0.42 |
| **Cancer** | 1 (0.6) | 0 | 1 (1.4) | 0.42 |
| **Transplant** | 1 (0.6) | 1 (1.05) | 0 | 1.0 |
| **Autoimmunity** | 1 (0.6) | 0 | 1 (1.4) | 0.42 |
| **BMI, med (ICC)** | 28.4 (25.3 – 32.7) | 27.6 (25.1 – 32.4) | 28.9 (25.1 – 32.8) | 0.45 |
| **Other drugs** |  |  |  |  |
| **Antibiotics** | 98 (59.3) | 57 (60) | 41 (58.5) | 0.85 |
| **Carbapenem drugs** | 68 (41.2) | 39 (41) | 29 (41.4) | 0.96 |
| **IBP** | 162 (98.1) | 93 (96.9) | 69 (98.5) | 1.0 |
| **Dexamethasone** | 134 (81.2) | 78 (82.1) | 56 (80) | 0.73 |
| **Ivermectin** | 105 (63.6) | 58 (60.4) | 47 (68.1) | 0.31 |

**Addendum 2.** Vital signs and oxygenation index on admission (Per-protocol analysis).

| **Variable** | **Entire Group (n=165)** | **Plasma (n=95)** | **IVIg (n=70)** | **P val** |
| --- | --- | --- | --- | --- |
| **MAP med (IQR)** | 93 (82 – 101) | 92 (82 – 100) | 95 (83 – 103) | 0.17 |
| **RF (IQR)** | 29 (24 – 34) | 30 (24 – 35) | 28 (24 – 32) | 0.26 |
| **O<sub>2</sub> Sat, med (IQR)** | 77 (66 – 85) | 78 (67 – 85) | 78 (67 – 85) | 0.91 |
| **PaO<sub>2</sub>/FiO<sub>2</sub>, med (IQR)** | 165 (96 – 240) | 166 (95 – 261) | 171 (110 – 242) | 0.88 |
| **PaO<sub>2</sub>, med (IQR)** | 55 (43 – 71) | 58 (45 – 77) | 53 (40 – 73) | 0.19 |
| **PaCO<sub>2</sub>, med (IQR)** | 29 (24 – 35) | 30 (25 – 35) | 28 (22 – 33) | 0.17 |

**Addendum 3.**
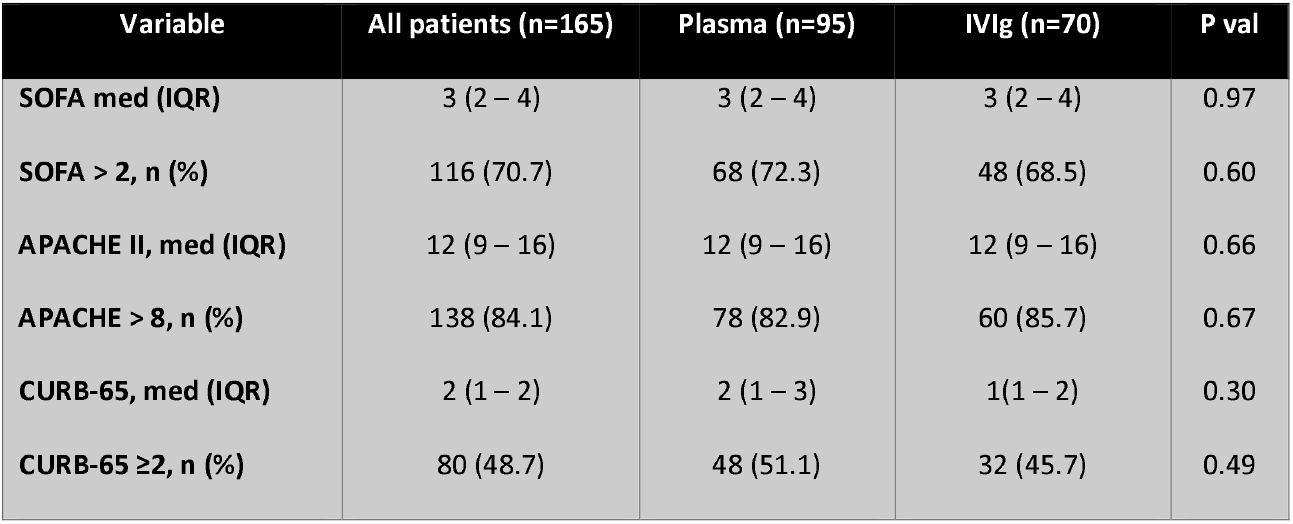
Prognostic Scales (Per-protocol analysis).

**Tabla 3.** Prognostic Scales (Intention-to-treat analysis).
| **Variable** | **All patients (n=165)** | **Plasma (n=95)** | **IVIg (n=70)** | **P val** |
| --- | --- | --- | --- | --- |
| **SOFA med (IQR)** | 3 (2 – 4) | 3 (2 – 4) | 3 (2 – 4) | 0.97 |
| **SOFA > 2, n (%)** | 116 (70.7) | 68 (72.3) | 48 (68.5) | 0.60 |
| **APACHE II, med (IQR)** | 12 (9 – 16) | 12 (9 – 16) | 12 (9 – 16) | 0.66 |
| **APACHE > 8, n (%)** | 138 (84.1) | 78 (82.9) | 60 (85.7) | 0.67 |
| **CURB-65, med (IQR)** | 2 (1 – 2) | 2 (1 – 3) | 1(1 – 2) | 0.30 |
| **CURB-65 ≥2, n (%)** | 80 (48.7) | 48 (51.1) | 32 (45.7) | 0.49 |

**Addendum 4.** Inflammation markers (Per-protocol analysis).

| Variable | All patients (n=165) | Plasma (n=95) | IVIg (n=70) | P val |
| --- | --- | --- | --- | --- |
| Med (IQR) |  |  |  |  |
| **Lactate** | 1.6 (1.1 – 2) | 1.5 (1.1 – 2.1) | 1.5 (1.1 – 2.1) | 0.84 |
| **Creatinine** | 0.9 (0.7 – 1.4) | 0.9 (0.7 – 1.4) | 1.0 (0.7 – 1.4) | 0.79 |
| **LDH** | 479 (367 – 658) | 464 (362 – 660) | 510 (385 – 658) | 0.26 |
| **Troponin** | 0.021 (0.01 -0.11) | 0.018 (0.01 – 0.06) | 0.018 (0.01 – 0.1) | 0.71 |
| **D-Dimer** | 1.3 (0.8 – 3.0) | 1.2 (0.7 – 3) | 1.3 ( 1 – 2.4) | 0.77 |
| **Ferritin** | 584 (324 – 1080) | 571 (268 – 1040) | 698 (358 – 1170) | 0.35 |
| **Hb** | 14.4 (13 – 15.5) | 14.2 (12.9 – 15.4) | 14.6 (13.6 – 15.7) | 0.05 |
| **Leukocytes** | 11.8(8.4 – 15.2) | 12 (8.7 – 16.2) | 11.8 (7.5 – 14.6) | 0.56 |
| **Neutrophils** | 9.9 (6.9 – 13.2) | 10.1 (6.9 – 14.2) | 10.1 (6.5 – 12.7) | 0.69 |
| **Lymphocytes** | 1.02 (0.7 – 1.5) | 1.06 (0.72 – 1.4) | 0.9 (0.7 – 1.4) | 0.59 |
| **CRP 1** | 192 (56 -270) | 98 (49 – 276) | 202 (66 – 265) | 0.66 |
| **CRP 2** | 163 (60 – 232) | 186 (62 – 235) | 86 (54 – 221) | 0.34 |
| **CRP 3** | 170 (47 – 231) | 157 (48 – 222) | 130 (38 – 204) | 0.83 |
| **Ferritin 1** | 584 (324 – 1080) | 571 (268 – 1040) | 598 (358 – 1170) | 0.35 |
| **Ferritin 2** | 710 (402 – 1280) | 747 (402 – 1300) | 645 (436 -1190) | 0.83 |
| **Ferritin 3** | 622 (342 – 1035) | 586 (337 -1040) | 687 (454 – 1210) | 0.74 |
| **D-Dimer 1** | 1.3 (0.8 – 2.9) | 1.2 (0.8 – 2.7) | 1.4 (1 – 2.4) | 0.50 |
| **D-Dimer 2** | 1.8 (1.1 – 4.3) | 1.5 (1.1 – 4) | 2.1 (1.3 – 4.1) | 0.18 |
| **D-Dimer 3** | 1.8 (0.9 – 2.5) | 1.6 (1.1 – 2.3) | 1.7 (0.8 – 3.2) | 0.80 |

**Addendum 5.** Complications (Intention-to-treat analysis and Per-protocol analysis). FIGURE LEGENDS

| **Variable, n (%)** | **All patients**<br>**(n=190)** | **Plasma (n=130)** | **IVIg (n=60)** | **P value** |
| --- | --- | --- | --- | --- |
| **Shock** | 136 (71.5) | 94 (72.3) | 42 (70) | 0.74 |
| **Myopathy** | 29 (15.2) | 20 (15.3) | 9 (15) | 0.94 |
| **Bacterial Infection** | 107 (56.6) | 75 (58.1) | 32 (53.3) | 0.63 |
| **Arrhythmias** | 9 (4.7) | 6 (4.2) | 3 (5) | 1.0 |
| **Heart Failure** | 3 (1.5) | 1 (0.77) | 2 (3.3) | 0.23 |
| **Myocardial Infarction** | 3 (1.5) | 3 (2.3) | 0 | 0.55 |
| **DKA** | 6 (3.1) | 5 (3.85) | 1 (1.6) | 0.66 |
| **Pressure Ulcers** | 13 (6.85) | 10 (7.6) | 3 (5.0) | 0.75 |
| **DVT** | 2 (1.05) | 2 (1.5) | 0 | 1.0 |
| **Pancreatitis** | 2 (1.05) | 2 (1.5) | 0 | 1.0 |
| **Hepatitis** | 1 (0.5) | 1 (0.7) | 0 | 1.0 |
| **Kidney Injury (KI)** | 56 (29.6) | 28 (29.2) | 18 (30.5) | 0.85 |
| **AKIN 1** | 19 (10) | 11 (8.4) | 8 (13.5) | 0.28 |
| **AKIN 2** | 9 (4.7) | 7 (5.3) | 2 (3.3) | 0.55 |
| **AKIN 3** | 28 (14.8) | 20 (15.3) | 8 (13.5) | 0.74 |

## Data Availability

Contact the corresponding author directly by email

